# Understanding depression in autism: The mediating role of mentalization, attachment, social support, and psychological flexibility

**DOI:** 10.1101/2024.07.12.24310324

**Authors:** Eszter Komoróczy, Dániel Sörnyei, Ágota Vass, Bálint Szuromi, Gabriella Vizin, János M. Réthelyi, Kinga Farkas

## Abstract

**Background:** Depression is highly prevalent in autistic individuals, yet its underlying psychological mechanisms remain insufficiently understood, especially in populations who self-identify as autistic but lack a formal diagnosis.

**Methods:** Using a moderated mediation approach (structural equation modeling), we investigated how autistic traits relate to depressive symptoms in three groups: individuals with clinically diagnosed autism (ASD, N = 136), self-identified autistic individuals without formal diagnosis (ASD-sd, N = 100), and neurotypical controls (N = 1566). Five psychosocial mediators were tested: mentalizing, anxious and avoidant attachment, perceived social support, and psychological flexibility.

**Results:** Depressive symptoms were significantly higher in both autistic groups compared to neurotypical controls. The association between autistic traits and depressive symptoms was fully mediated by a combination of psychological flexibility, attachment style, mentalizing, and perceived social support. Among these, psychological flexibility emerged as the strongest and most consistent mediator across all groups. In contrast, mentalizing showed an indirect effect only in the non-ASD group, while avoidant attachment was a significant mediator solely in the clinically diagnosed ASD group. Perceived social support showed a modest indirect effect only in the non-ASD group.

**Conclusions:** Our findings support a transdiagnostic model in which psychological flexibility plays a central protective role against depression across varying levels of autistic traits. Meanwhile, attachment-related and social-cognitive vulnerabilities contribute in different ways depending on diagnostic status. These results underscore the clinical importance of individualized interventions targeting such modifiable factors. Mentalization-based treatment, adapted for neurodiverse populations, may offer a promising framework for addressing both common and subgroup-specific mechanisms.

**Lay abstract:** Many autistic people experience depression, but we still know little about why this happens and what psychological factors are involved, especially for those who see themselves as autistic but don’t have a formal diagnosis. We studied three groups: people with diagnosed autism, people who self-identify as autistic, and people without autism. We wanted to understand which psychological traits might explain why autistic traits are linked to depression. We found that a skill called psychological flexibility (being able to stay present and adapt to difficult emotions) was the most important protective factor in all groups. Other factors, like how people think about relationships (attachment) and how well they understand themselves and others (mentalizing), were only important for some groups. These findings suggest that therapies which improve flexibility and help with emotional connection may reduce depression in autistic people and those with high autistic traits.

## Introduction

### What is the relevance of studying depression in autism?

Autism spectrum disorder (ASD) is a neurodevelopmental condition characterised by difficulties in social communication and interaction, restricted or repetitive behaviours or interests, and sensory processing alterations (American Psychiatric Association, 2013, 2022). The rising prevalence of ASD (Zeidan et al., 2022) makes it increasingly crucial to understand its comorbidities, particularly the frequent and severe co-occurrence of depression (Bougeard et al., 2021). A number of studies have examined the relationship between ASD and depression, suggesting that individuals with autism have a higher rate of depression than the population average (e.g. (Hollocks et al., 2019; Lai et al., 2019; Wigham et al., 2017). While the prevalence of depressive disorders is estimated between 7-11% in the general population (Lim et al., 2018; Wittchen et al., 2011; Wittchen & Jacobi, 2005), it is approximately 10-15%, ranging from 2.5% to 47.1% in ASD (Hossain et al., 2020), and individuals with ASD are four times more likely to experience depression during their lifetime compared to neurotypical individuals (Hudson et al., 2019) according to recent meta-analyses. Despite the high prevalence of depressive symptoms in ASD patients, the nature of the relationship between the two conditions remains uncertain.

As depression is a multifactorial disorder and ASD is a heterogeneous spectrum condition, it is difficult to single out one causal factor. It has been recognised that one in two individuals with autism has alexithymia (Albantakis et al., 2020); moreover loneliness (Schiltz et al., 2021; Umagami et al., 2022), emotion regulation deficits, and intolerance of uncertainty have also been found to be associated with symptoms of anxiety and depression in autistic populations (Cai et al., 2018). Furthermore, depressive symptoms severely impact the quality of life (Thiel et al., 2024).

Although autism spectrum disorder (ASD) is typically diagnosed by professionals using standardized criteria (e.g., DSM-5, ICD-11), an increasing number of individuals self-identify as autistic, particularly those who may have been overlooked by traditional diagnostic pathways, such as women and gender-diverse individuals who often present atypical or camouflaged symptoms (Hull et al., 2017; Lai & Baron-Cohen, 2015). These self-diagnosed individuals frequently report experiences consistent with the autistic phenotype and may experience comparable psychological distress (Banker et al., 2025; McDonald, 2020). Thus, their inclusion may provide a more comprehensive understanding of the autistic trait continuum and its association with depression.

In this article, we explore the interconnected phenomena of mentalization, attachment, peer support and psychological flexibility as potential mediators to understand their combined impact on the relationship between ASD and depressive symptoms. Our inquiry aligns with the mentalization theory proposed by Fonagy, Luyten, and Bateman, subsequently extended to illuminate diverse psychopathological phenomena (Fonagy & Bateman, 2016; Fonagy & Luyten, 2009; Luyten et al., 2020). By this exploration, we endeavour to shed light on the intricate web connecting ASD and depressive symptomatology. In line with this spectrum perspective, our study includes individuals with both formal and self-reported ASD diagnoses, allowing us to capture a broader range of autistic experiences. This is particularly relevant given that many self-identified autistic individuals exhibit elevated autistic traits and similar patterns of psychological vulnerability (Banker et al., 2025), even in the absence of formal diagnosis.

### What is the role of mediating factors in the relationship between autism and depression?

#### Mentalization

Social communication deficits are core symptoms of ASD. Mentalizing deficits occur early but remain observable throughout life (Baron-Cohen, 2000; Chung et al., 2014). Research during recent years in ASD has consistently shown deficits in theory of mind (ToM; one specific aspect of mentalization) (Yirmiya et al., 1998), emotion perception and processing (Brewer et al., 2017; Velikonja et al., 2019), empathy, affective mentalizing, and cognitive mentalizing and related altered neural functioning (Arioli et al., 2021). ASD is, however, a heterogeneous spectrum; it is important to note that within the condition, variability in mentalizing abilities is not negligible (Lombardo et al., 2016).

Mentalization deficits are associated with depressive symptoms as well. Identifying and interpreting mental states of the self and others have been found to be impaired among patients with depression, and the degree of mentalizing impairment was associated with the severity of depression and worse clinical outcomes (Fischer-Kern et al., 2013; L. Lee et al., 2005; Lombardo et al., 2016). Deficits in theory of mind were shown to be a predictor of relapse and impairment in social functioning in depression (Inoue et al., 2006) and were associated with cognitive deficits as well (Wang et al., 2008). Fischer-Kern et al. (2022) also showed that, besides mentalizing deficits, attachment insecurity and unresolved loss are also important predictors of depression. A current integrative model of depression, the stress-reward-mentalizing model suggests a developmental cascade. This sequence involves elevated stress levels and persistent heightened arousal, potentially coupled with genetic predisposition, culminating in impaired reward sensitivity within attachment relationships and the realm of agency and self-governance. Consequently, this progression impacts the capacity for mentalizing and social cognitive functions (Luyten & Fonagy, 2018).

#### Attachment

Emotion recognition, reciprocal social communication, ToM, and other aspects of mentalization are essential to form secure attachment and maintain a balanced parent-child relationship (Fonagy et al., 1991; Keenan et al., 2017). Children with ASD show impairments in social, and emotional capacities from an early age (American Psychiatric Association, 2022; Baron-Cohen et al., 1985; Cassel et al., 2007; Dawson et al., 2004; Nuske et al., 2013). They exhibit heightened attachment insecurity (Naber et al., 2007), as well as decreased sensitivity in their relationships with caregivers (Rutgers et al., 2004; Van IJzendoorn et al., 2007), including more contact resistance and less contact-seeking behaviours (Rogers et al., 1993), fewer pro-social responses to caregivers and difficulty being soothed (Grzadzinski et al., 2014), and less frequent looking, showing, smiling, and mutual play behaviours (Akdemir et al., 2009; Dissanayake & Crossley, 1997). According to a recent review, only 47% of children with ASD demonstrated secure attachments (Teague et al., 2017). The notion that individuals with ASD have difficulties to form attachments, however, is tempered by the observation that they express a preference for the caregiver over a stranger, and experience distress when unexpectedly separated from them (Capps et al., 1994; Dissanayake & Crossley, 1997; Koren-Karie et al., 2009).

Employing questionnaire-based measures, instead of observation-based paradigms like the Strange Situation (Ainsworth et al., 1978), during middle childhood, no differences in attachment security were found between autistic and neurotypical participants (Bauminger et al., 2010; Chandler & Dissanayake, 2014; Wu et al., 2015). Upon reaching adulthood, attachment style remains relatively stable (Fraley, 2002), but the object of attachment shifts from parents to peers, including friends and romantic partners (Doherty & Feeney, 2004; Hazan & Zeifman, 1994; Nickerson & Nagle, 2005). Insecure attachment, especially avoidant attachment style, is more prevalent in ASD groups than in the general population (Taylor et al., 2008), and associated with autistic traits in non-clinical samples (Gallitto & Leth-Steensen, 2015), and it is associated with lower relationship satisfaction (Beffel et al., 2021). A recent study found that adult attachment styles are important predictors of mental health: higher anxious attachment predicted higher depression and anxiety, interestingly, higher avoidant attachment predicted lower anxiety and stress in autistic adults (Lee et al., 2022).

Attachment-related distress and avoidance have been associated with psychopathology (Mikulincer & Shaver, 2012, 2017), including anxiety (Bosmans et al., 2010) and depression (Beatson & Taryan, 2003; Besser & Priel, 2003; Cantazaro & Wei, 2010; Dagan et al., 2018; Malik et al., 2015). Individuals reporting low attachment anxiety and avoidance have the ability to maintain interpersonal relationships through secure attachment and report higher levels of adaptive emotion regulation, positive self- and other perceptions, and greater psychological well-being (Mikulincer & Shaver, 2019; Raque-Bogdan et al., 2011). Attachment style has an impact on emotional, behavioural, and relational patterns (Mikulincer & Shaver, 2017), resilience to stressful life events (Mikulincer & Shaver, 2012), and the quality of interpersonal relationships (Collins, 1996; Shaver et al., 2000; Torquati & Vazsonyi, 1999).

#### Social support

Developmental research on mentalization suggests that environmental influences play a significant role in the development of mentalization. In recent years, there has been a notable shift in this respect: while earlier theories (Fonagy et al., 1991) emphasised the specific role of dyadic (parent-child) attachment in facilitating or inhibiting the development of mentalization, recent views have taken a more comprehensive approach. Accordingly, in addition to the family environment, peers, and wider sociocultural factors are in close interaction with attachment, mentalization and social learning, through the capacity to rely on others to provide social information (epistemic trust) and the ability to benefit from the positive effects of the environment (salutogenesis) (Fonagy & Allison, 2014; Luyten et al., 2020). As ASD is linked to difficulties in social communication and interaction, people with ASD are less able to form social relationships and access and incorporate the benefits of such peer support. The effect of perceived social support has been measured mainly among the parents of children with ASD (Alon, 2019; He et al., 2022; Lu et al., 2021). Studies examining social support in adult ASD found that social support was associated with psychological, social, and environmental well-being (Bishop-Fitzpatrick et al., 2018; Leader et al., 2021; Tsermentseli, 2022).

Social support is a protective factor against depression, with parental support as the most influential during childhood and adolescence, whereas spouses, family, and friends become more important in adulthood and senior age (Gariépy et al., 2016). Perceived social support mediated loneliness and depression in the elderly (Liu et al., 2016), lower quality support predicted a sixfold risk of depressive symptoms among college students (Hefner & Eisenberg, 2009), and elevated the risk of postpartum depression (Akbari et al., 2020), and more importantly showed to be a moderator between negative life events and depression in adolescence (Miloseva et al., 2017). On the contrary, higher levels of perceived social support were linked to positive affect, overall life satisfaction, and favourable outcomes of mental disorders (Siedlecki et al., 2014; J. Wang et al., 2018).

#### Psychological flexibility and ASD

Psychological resilience and flexibility are also important factors in mental health (Kashdan & Rottenberg, 2010; Rutten et al., 2013; Yasinski et al., 2020). Psychological inflexibility is crucial in understanding the relationship between depression and autism due to its effects on coping strategies, emotional regulation, and adaptive functioning. Individuals with autism often struggle to adapt to changing circumstances. Inflexibility can appear as rigid thinking patterns, resistance to change, and difficulty shifting focus from negative thoughts or experiences, which can intensify their depressive symptoms. Although ASD is characterised by behavioural inflexibility (American Psychiatric Association, 2022), the role of psychological inflexibility was measured only once among autistic adults. A recent study reported that psychological flexibility (measured by AAQ-II) and attachment anxiety mediated the association between childhood experiences of parenting and mental health in later life in individuals with autism (Lee et al., 2022). It is conceivable that psychological flexibility can be instrumental in several ways when considering its reciprocal relationship with attachment, mentalizing, and perceived social support in individuals with autism. It can also facilitate considering alternative perspectives, managing conflicting emotions, adapting their responses in interpersonal challenges, and being open to seeking and receiving support.

### Hypotheses and aims of the current paper

In this study, we aim to validate the heightened prevalence of depression in individuals with autistic traits through a comparative analysis of depressive symptom severity. First, we examine the associations between autistic traits and depressive symptoms across both neurotypical and autistic populations. Then, we investigate whether this relationship is mediated by key psychological constructs: mentalization, attachment style, perceived social support, and psychological flexibility - factors that are known to influence mental well-being in both autism and the general population. Finally, we explore whether these pathways differ based on diagnostic status, including both individuals with a formal ASD diagnosis and those who self-identify as autistic. By including the self-diagnosed group, we aim to determine whether their psychological profiles align more closely with clinically diagnosed individuals or represent a distinct subgroup along the autism spectrum.

## Methods

### Recruitment procedure, ethical concerns

Participants were requested to complete an online survey between September 2021 and April 2022. Initially, we reached out to outpatient units, institutions, and counsellors specialising in aiding individuals diagnosed with autism spectrum disorder, with the purpose of facilitating questionnaire accessibility for those with an established ASD diagnosis.

The language of the questionnaires was Hungarian and the wording was designed for maximum clarity. Experts from these organisations also checked the questionnaire, provided suggestions and these were incorporated in the final version of the survey. Subsequently, we also recruited non-ASD participants through media and social media advertisements.

Participants provided informed consent online. The authors assert that all procedures contributing to this work comply with the ethical standards of the relevant national and institutional committees on human experimentation and with the Helsinki Declaration. The study was approved by the Semmelweis University Regional and Institutional Committee of Science and Research Ethics, SE RKEB: 159/2021.

### Participants

Overall 2409 participants were recruited and started the survey. Completing the whole survey took about 40 minutes, and 1783 participants answered all the items. Participants were allowed to stop at any point and resume filling out the questionnaires later. We have constructed four selection criteria to ensure high quality of our data. First, we have excluded those participants who did not fill out the BDI, or the AQ questionnaires which were intended to measure the depressive and autistic symptom severity, respectively. Participants between the ages of 18 and 65 were included. Further, we have confirmed that all of our participants have completed at least 8 years of elementary education, or have provided realistic data regarding the years they had spent in education: we have excluded all participants who claimed to have spent more years in education than their age minus 5 years. Finally, we have also rigorously screened our sample based on the time spent filling out the questionnaires that were of most interest to us. As participants were allowed to take a break and resume filling out the questionnaires later at their convenience, there were some extremely high values in the time spent filling out many of the questionnaires. Thus, we decided to assess the time it took participants to fill out the respective questionnaires by calculating measures that are more robust to outliers. We have calculated the median filling time and median absolute deviation of filling time as a measure of spread for each questionnaire. We have excluded those participants who have spent less than the median minus one and a half median absolute deviation time on the following questionnaires: AQ, BDI, MZQ, AAS, MSPSS, and AAQ-II.

### Questionnaires

#### Demographic data collection

Demographic information was collected using a structured questionnaire administered at the beginning of the study. Participants were asked about their age, gender, education level, socioeconomic and marital status.

#### Autism Spectrum Quotient (AQ)

Autism Spectrum Quotient is a 50-item self-report questionnaire to measure the degree of autistic traits in adults with normal intelligence. The AQ assesses five different areas: social skill, attention switching, attention to detail, communication, and imagination; 10 items cover each area, on a 4-point Likert scale with values of ‘definitely agree’, ‘slightly agree’, ‘slightly disagree’ or ‘definitely disagree’. At the end of the questionnaire, each participant was given feedback on their score on the AQ scale and whether they should seek professional advice. The internal consistency of items in each of the five domains and Cronbach’s alpha coefficients were all moderate to high (Baron-Cohen et al., 2001). In the present study, the overall reliability was good (total ⍺ = 0.89, ASD ⍺ = 0.86, ASD-sd ⍺ = 0.85 and NTP (non-ASD) ⍺ = 0.86).

*Beck Depression Inventory (BDI)* is a widely used 21- item, self-report inventory measuring symptoms of depression with good internal consistency (⍺ = 0.86) (Beck et al., 1988). In our study, this scale had good internal consistency (total ⍺ = 0.90, ASD ⍺ = 0.91, ASD-sd ⍺ = 0.88 and N T P ⍺ = 0.90).

#### Mentalization Questionnaire (MZQ)

The Mentalization Questionnaire is a 15-item self-report measure of mentalization (Hausberg et al., 2012). The items of the MZQ cover four domains of mentalization: ‘Refusing self-reflection’, ‘Emotional awareness’, ‘Psychic equivalence mode’, and ‘Regulation of affect’. The structural validity of the Hungarian version of the MZQ was assessed among psychotic patients and the study confirmed the four-factor structure which differs from healthy samples (Fekete et al., 2019). In our study, however, we used the overall MZQ score (higher scores meant greater difficulty in mentalisation), which had good reliability in the sample (total ⍺ = 0.86, ASD ⍺ = 0.82, ASD-sd ⍺ = 0.77 and N T P ⍺ = 0.85).

#### Adult attachment scale (AAS)

Adult attachment style was measured by the Adult Attachment Scale (AAS), which consists of 18 Likert items that can be grouped into two subscales (Collins, 1996). Psychometric properties of the AAS were evaluated in a Hungarian sample recently (Őri et al., 2021). In our study, we used the scores reflecting anxious and avoidant attachment styles, which had good internal consistency (total ⍺ *(anxious)* = 0.88 and ⍺ *(avoidant)* = 0.86, ASD ⍺ *(anxious)* = 0.87 and ⍺ *(avoidant)* = 0.83, ASD - sd ⍺ *(anxious)* = 0.88 and ⍺ *(avoidant)* = 0.81, and N T P ⍺ *(anxious)* = 0.88 and ⍺ *(avoidant)* = 0.86).

#### Multidimensional Scale of Perceived Social Support (MSPSS) during adolescence

Social support during adolescence was measured with the Multidimensional Scale of Perceived Social Support (MSPSS), which has 12 Likert items (Canty-Mitchell & Zimet, 2000; Papp-Zipernovszky et al., 2017). Participants were asked to focus on their adolescence. In our study, this scale had excellent internal consistency (total ⍺ = 0.95, ASD ⍺ = 0.92, ASD-sd ⍺ = 0.90 and N T P ⍺ = 0.95).

#### Acceptance and Action Questionnaire-II (AAQ-II)

AAQ - II is a measure of psychological flexibility, which has 7 Likert items (7-point scale, from “never true” to “always true”). Higher total scores mean less flexibility. The tool has satisfactory psychometric properties (⍺ = 0.84) (Bond et al., 2011), and in a Hungarian sample the internal consistency was good (⍺ = 0.90) (Eisenbeck & Szabó-Bartha, 2018). In our study, this scale had good internal consistency (total ⍺ = 0.93, ASD ⍺ = 0.92, ASD-sd ⍺ = 0.90 and NTP ⍺ = 0.93).

### Data Analysis

R, RStudio 2024.09.1 software was used to perform the analyses and data visualization. First, the Krukskal-Wallis, Wilcoxon rank sum test and Fisher’s exact test statistics were used to compare the ASD and non-ASD samples (Tables 1 and 3). Second, receiver operating characteristic (ROC) curve analysis was performed to examine the sensitivity and specificity of the AQ to determine a scoring cutoff value that discriminates between autistic and non-autistic cases (Hosmer, 2013; Robin et al., 2011). In the third step, we conducted Spearman correlation analyses to explore the associations between AQ scores and other psychological constructs. We compared the correlation coefficients between ASD and non-ASD groups by z-test (Eid et al., 2013). Detailed results and methods are reported in the Supplementary Material (Figure S5).

**Table 1.**
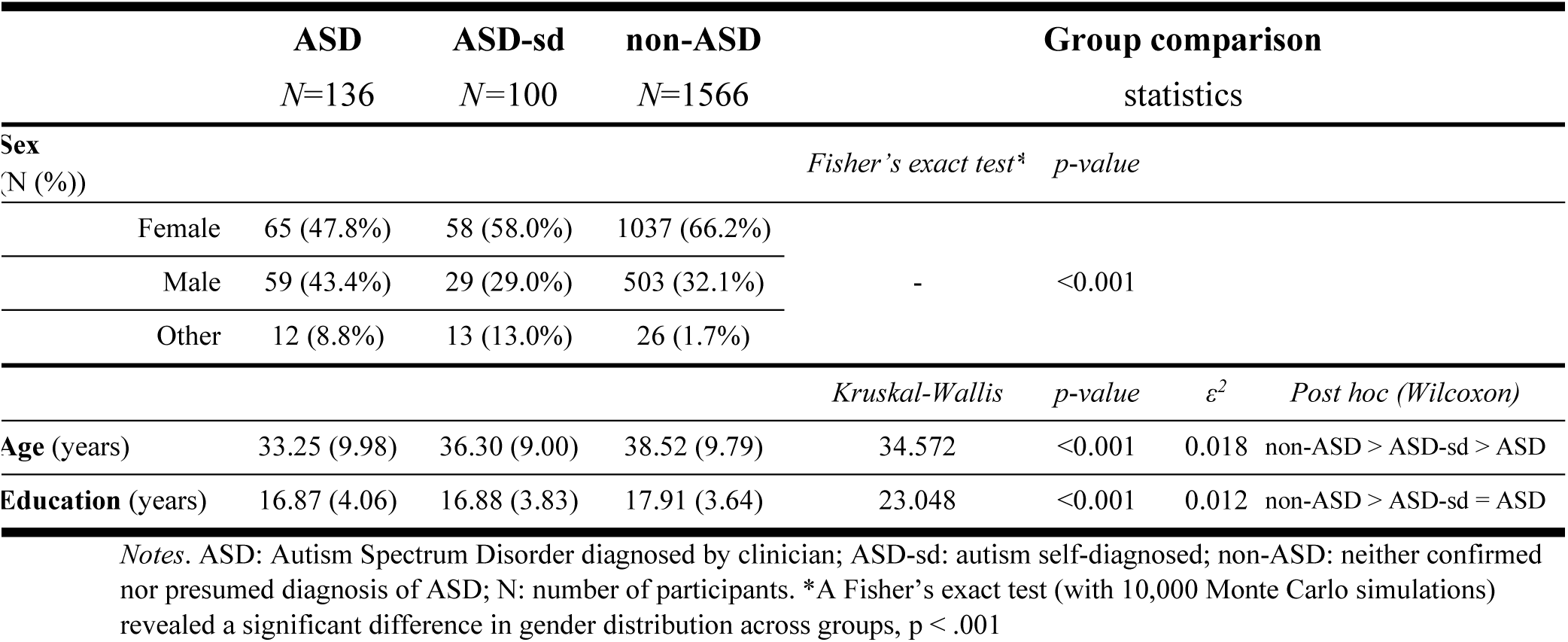
Basic demographics.

**Table 3.** Clinical characteristics – questionnaire scores.

| *Questionnaire* | **ASD**<br>*N*=133 |  |  | **ASD-sd**<br>*N*=100 |  |  | **non-ASD**<br>*N*=1550 |  |  |
| --- | --- | --- | --- | --- | --- | --- | --- | --- | --- |
|  | *Mean* | *Median* | *SD* | *Mean* | *Median* | *SD* | *Mean* | *Median*<br>*n* | *SD* |
| AQ sum | 32.27 | 34.0 | 8.16 | 32.82 | 34.0 | 7.22 | 20.29 | 20.0 | 8.21 |
| social | 6.79 | 8.0 | 2.64 | 6.80 | 7.0 | 2.31 | 3.82 | 3.0 | 2.81 |
| attention switch | 7.91 | 8.0 | 2.02 | 8.01 | 8.5 | 2.01 | 5.20 | 5.0 | 2.50 |
| communication | 6.42 | 6.0 | 2.50 | 6.38 | 6.0 | 1.94 | 3.03 | 3.0 | 2.20 |
| attention to detail | 6.35 | 7.0 | 2.02 | 6.65 | 7.0 | 2.20 | 5.10 | 5.0 | 2.14 |
| imagination | 4.81 | 4.5 | 2.22 | 4.98 | 5.0 | 2.19 | 3.13 | 3.0 | 1.95 |
| BDI | 16.59 | 15.5 | 11.53 | 17.08 | 17.0 | 9.91 | 11.74 | 10.0 | 8.86 |
| MZQ | 36.60 | 38.0 | 10.70 | 37.01 | 37.0 | 9.10 | 25.78 | 26.0 | 11.03 |
| AAS anxious | 20.79 | 21.0 | 6.55 | 20.48 | 21.0 | 6.82 | 16.16 | 16.0 | 6.46 |
| AAS avoidant | 40.75 | 41.5 | 10.03 | 43.66 | 45.0 | 8.63 | 34.95 | 35.0 | 9.45 |
| MSPSS | 35.10 | 36.0 | 11.62 | 32.95 | 31.0 | 9.87 | 41.72 | 43.0 | 11.87 |
| AAQ-II | 31.97 | 32.0 | 9.88 | 31.66 | 32.0 | 9.23 | 22.84 | 22.0 | 10.05 |

Notes. ASD: Autism Spectrum Disorder diagnosed by clinician; non-ASD: neither confirmed nor presumed diagnosis of ASD; N: number of participants. SD: standard deviation. AQ: Autism Spectrum Quotient; BDI: Beck Depression Inventory; AAS anxious: Adult Attachment Style: Anxious attachment; AAS avoidant: Adult Attachment Style: Avoidant attachment; MZQ: Mentalization Questionnaire; MSPSS: Multidimensional Scale of Perceived Social Support; AAQ-II: Acceptance and Action Questionnaire-II

### Moderated mediation

Finally, to investigate the conditional indirect effects of autistic traits on depressive symptoms, we conducted multivariate moderated mediation analyses using structural equation modeling (SEM) with the *lavaan* package (version 0.6-19) in R (Rosseel, 2012). This approach implements regression-based moderated mediation analysis by modeling all mediators simultaneously within a unified SEM framework. Interaction terms between the predictor (AQ total score) and diagnostic groups (ASD, ASD-sd as dummies) were included to estimate group-specific a- paths (predictor → mediator) and b- paths (mediator →outcome). Attachment style (AAS scores), mentalization (MZQ total score), perceived social support (MSPSS total score), and psychological flexibility (AAQ - II total score) had been set as mediators and depression severity index (BDI score) was the outcome variable. To control for potential confounding, sex (three categories: male, female, other), age (continuous), and years of education (continuous) were included as covariates predicting both the mediators and the outcome. Standardized estimates are reported. Indirect effects were calculated for each group separately, and significance was determined via bias-corrected bootstrap confidence intervals based on 5000 resamples. This method enabled robust estimation of complex, group-differentiated mediation pathways, accounting for shared variance among mediators and allowing direct comparison of conditional effects across groups. Model fit was evaluated using multiple indices (CFI, T LI, RMSEA, SRMR), although the complexity and moderation structure were expected to lower absolute fit indices (Supplementary Material T able S1)

### Sensitivity analysis

To explore whether self-diagnosed and clinically diagnosed individuals could be merged analytically, we also tested models combining both groups (Supplementary material, Sensitivity analysis, Figures S6-7, Tables S7-10).

## Results

### Descriptive statistics

The basic demographic data of the participants are shown in Table 1. The gender distribution in the non-ASD group reflects the generally higher female response rate (female: 66.2% vs. male: 32.1%), 1.7% of respondents identified themselves as other gender. In the ASD group, by contrast, the proportion of women and men showed a nearly even balance (female: 47.8% vs. male: 43.4%; as men are more likely to be diagnosed with ASD, but women tend to be more willing to answer questionnaires), and there was a significantly higher proportion of those who defined themselves as other gender (8.8%). For completeness, we also received responses from individuals who believed they were on the autism spectrum or were perceived as such by friends or relatives, but who did not have a formal diagnosis at the time of completing the survey. Their AQ scores (See Supplementary material Figure S1-S4 & ROC analysis) suggest a real possibility that their assumption is correct; however, it was not possible to formally investigate or diagnose them. Sensitivity analysis yielded qualitatively similar results, suggesting robustness, but group-specific analyses revealed important nuanced psychological differences, justifying the use of separate groups in the main model. Basic demographic data are shown in Table 1, while detailed demographic data for the three study groups are presented in Supplementary Material Table S1. As there were significant differences in sex, age, and education level across the groups, these variables were included as covariates in subsequent analyses.

### Clinical characteristics

The ASD and non-ASD groups scored differently on all questionnaires, with the ASD group scoring significantly higher on AQ, BDI, MZQ, AAS (both insecure attachment subscale), and AAQ-II, and lower on the MSPSS scale, the latter indicates lower perceived peer and family support during adolescence. Detailed data and between-group differences are shown in Table 3 and Figure 1.

**Figure 1.**
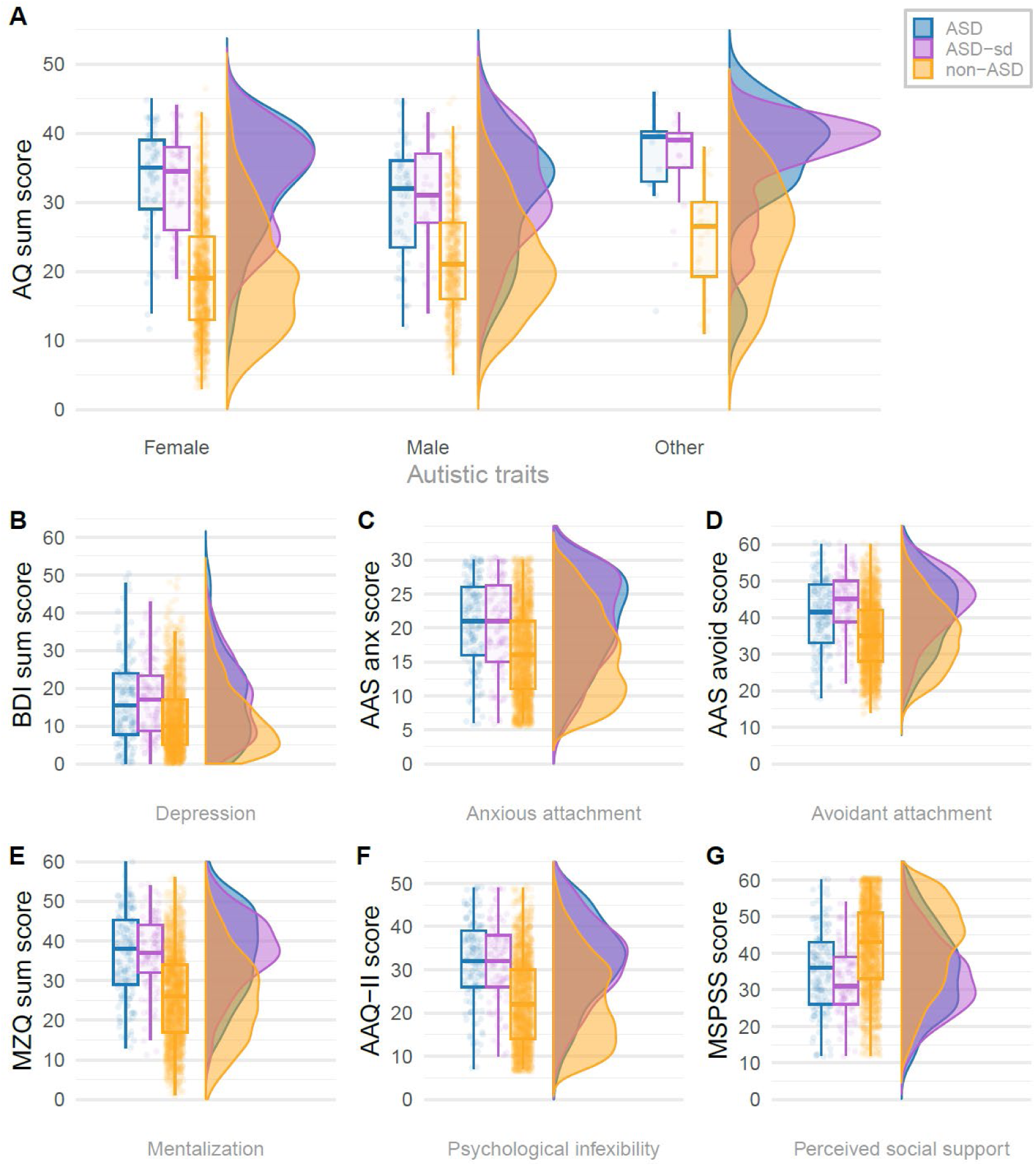
Distribution of clinical characteristics (questionnaire scores) by group. *Notes*: Blue: participants diagnosed with ASD; Purple: participants with self-diagnosed autism spectrum disorder; Yellow: participants with neither confirmed nor presumed diagnosis of ASD; Boxplot: the box shows the interquartile range, the horizontal line indicates the median, and the whiskers show the maximum and minimum values. A) The y axis shows the AQ (Autism Spectrum Quotient) scores of participants. On the x axis participants are assigned into three groups (participants diagnosed with ASD and participants not diagnosed with ASD and are neither self-diagnosed) and participants are further grouped based on their sex. B-G) The y axis shows the further scores of participants. On the x axis participants are assigned into three groups. BDI: Beck Depression Inventory; AAS anxious: Adult Attachment Style: Anxious attachment; AAS avoidant: Adult Attachment Style: Avoidant attachment; MZQ: Mentalization Questionnaire; MSPSS: Multidimensional Scale of Perceived Social Support; AAQ-II: Acceptance and Action Questionnaire-II

In Figure 1A AQ scores were also plotted by both diagnostic group and sex, revealing patterns similar to those reported by Baron-Cohen et al. (2014), showing an attenuation of typical sex differences in the autism group.

### Group differences in trait measures

Statistically significant group differences were found across all questionnaire measures (Kruskal–Wallis tests, all *p*s < .001). Post-hoc comparisons revealed that both individuals with clinically diagnosed ASD and those with self-diagnosed ASD scored significantly higher than non-ASD participants on all measures of autistic traits, depression, psychological inflexibility, attachment insecurity, and mentalization difficulties (all adjusted *p*s < .001). A similar pattern was observed in the opposite direction for perceived social support, with lower scores among ASD groups, consistent with its role as a protective factor. Differences between the ASD and ASD-sd groups were generally non-significant, except for avoidant attachment (AAS-avoid), where the ASD-sd group reported higher scores (adjusted *p* = .025, *r* = .15). For detailed test statistics, effect sizes, and adjusted *p*-values, see Supplementary Table S2.

### Sensitivity and specificity of the AQ

ROC analysis indicates that AQ-50 scores effectively distinguish both clinically diagnosed and self-diagnosed autistic individuals from non-ASD participants, with AUCs of 0.842 and 0.865, respectively. While the self-diagnosed group showed slightly higher discrimination, a DeLong test revealed that this difference was not statistically significant (*D* = –0.924, *p* = .355), indicating comparable classification performance across groups. The optimal cutoff score was higher for the ASD (clinically diagnosed) group (29.5) than for the self-diagnosed group (24.5). Sensitivity was higher in the diagnosed group (0.844 vs. specificity: 0.699), whereas specificity was higher in the self-diagnosed group (0.86 vs. sensitivity: 0.699), suggesting that self-identified individuals show meaningful autistic traits but may present a more heterogeneous or subclinical profile (see details in the Supplementary material Figures S1-S4). Accordingly, we included both groups separately in the main *lavaan* SEM and group comparisons to test whether self-diagnosed individuals represent a meaningful subtype or a distinct point along the autistic trait continuum. To ensure the robustness of our findings, we also report a sensitivity analysis using a combined ASD group (clinically diagnosed and self-diagnosed) in the Supplementary Material (Figures S6-7, Tables S7-S10). This yielded qualitatively similar results, and model fit did not change significantly. Therefore, in line with our exploratory aims, we retained the three-group structure in the main analyses to examine potential nuanced psychological differences among the study groups.

### Moderated mediation model

A moderated mediation model was estimated using structural equation modeling (SEM) in *lavaan*, controlling for age, years of education, and sex. The model fit was poor (χ²(60) = 3441.42, *p* < .001, CFI = 0.58, TLI = 0.14, RMSEA = 0.181, SRMR = 0.056), but the individual pathways were interpreted according to the pre-specified hypotheses. Indirect effects were calculated for each mediator across the three groups (non-ASD, ASD, ASD-sd). Mediation analyses revealed that psychological inflexibility (AAQ-II) significantly mediated the association between autistic traits (AQ) and depressive symptoms (BDI) across all groups. In the non-ASD group the indirect effect was *β* = 0.267, SE = 0.020, 95% CI [0.229, 0.308], *p* < .001, in the ASD group: *β* = 0.121, SE = 0.058, 95% CI [0.026, 0.250], *p* = .035 and in ASD-sd group *β* = 0.144, SE = 0.069, 95% CI [0.024, 0.290], *p* = .037. Avoidant attachment (AAS-avoid) significantly mediated the AQ → BDI relationship only in the clinically diagnosed ASD group: the indirect effect was *β* = 0.115, SE = 0.051, 95% CI [0.026, 0.222], *p* = .025. Indirect effects through AAS-avoidance were smaller and non-significant in the non-ASD (indirect effect: *β* = 0.020, SE = 0.020, 95% CI [-0.019, 0.059], *p* = .314) and ASD-sd groups (indirect effect: *β* = 0.111, SE = 0.070, 95% CI [-0.009, 0.266], *p* = .113). Mentalization (MZQ) showed a significant indirect effect only in the non-ASD group (*β* = 0.115, SE = 0.021, 95% CI [0.074, 0.157], *p* < .001). The mediation paths via perceived social support (MSPSS) were weak and non-significant in ASD and ASD-sd groups; a marginal effect was observed in the non-ASD group (*β* = 0.022, SE = 0.011, 95% CI [–0.0002, 0.044], *p* = .054). Anxious attachment (AAS-anx) did not show significant mediation in any group. (Figure 2, Indirect mediators). These effects remained consistent after controlling for age, sex, and education. Detailed parameter estimates, confidence intervals are provided in the Supplementary Material (Tables S2–S3).

**Figure 2.**
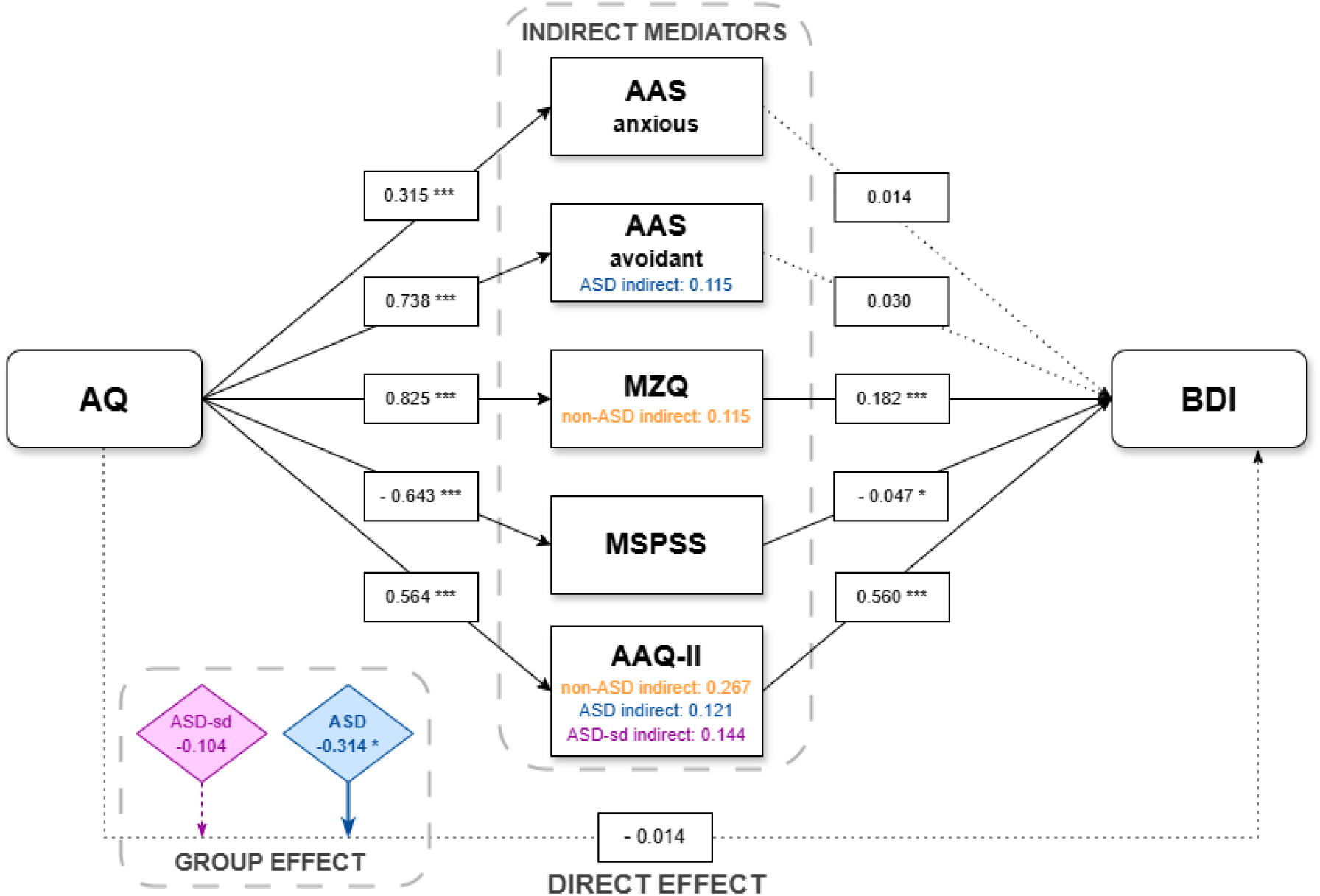
Moderated mediation model. *Notes.* Blue: participants diagnosed with ASD; purple: participants with self-diagnosed autism spectrum disorder; yellow: participants with neither confirmed nor presumed diagnosis of ASD. Predictor: AQ = Autism Spectrum Quotient; outcome: BDI = Beck Depression Inventory; mediators: AAS anxious = Adult Attachment Scale: attachment anxiety; AAS avoidant = Adult Attachment Scale: attachment avoidance; MZQ = Mentalization Questionnaire; MSPSS = Multidimensional Scale of Perceived Social Support; AAQ-II = Acceptance and Action Questionnaire-II. Standardised regression coefficients (β) are shown on the arrows. Continuous lines indicate significant, and dotted lines indicate non-significant regression effects. Levels of significance: * *p* < .05; ** *p* < .01; *** *p* < .001

### Direct Effect

The direct effect of AQ on BDI was nonsignificant when controlling for mediators and covariates (standardized estimate: *β* = -0.014, SE = 0.028, 95% CI [-0.068, 0.041], *p* = .619) (Figure 2, Direct effect). However, group membership significantly predicted depression: individuals in the ASD group had higher depression scores compared to the non-ASD group (*β* = –0.314, *p* = .038). No significant direct effects were found for the ASD-self-diagnosed group (*p* = .499) (Figure 2, Group effect) or for the interaction terms between AQ and group status (all *p*s > .4), for details see Supplementary Material Table S2. These results suggest that the association between autistic traits and depression was primarily indirect; psychological inflexibility is the primary mediator linking autistic traits to depressive symptoms across groups, avoidant attachment plays a specific mediating role in individuals with ASD, while mentalization in non-ASD.

### Group Comparisons of Indirect Effects

To visualize how the path estimates vary across diagnostic groups, we plotted the a-paths, b-paths, and resulting indirect effects for each mediator (see Figure 3).

**Figure 3.**
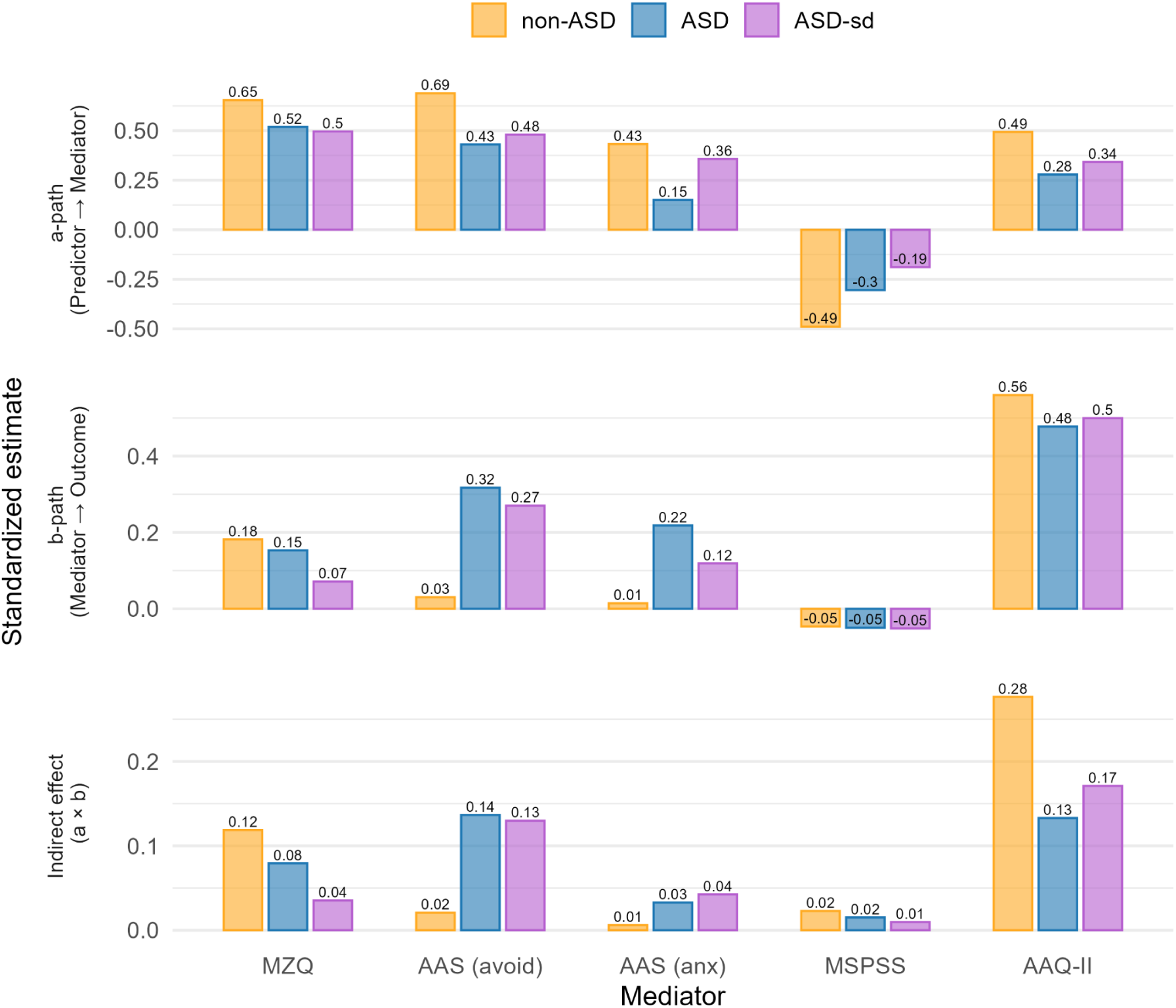
Standardized estimates of a-paths, b-paths, and indirect effects (a × b) across diagnostic groups for each mediator. *Notes*. Values are based on SEM estimates adjusted for age, sex, and education. Blue: participants diagnosed with ASD; purple: participants with self-diagnosed autism spectrum disorder; yellow: participants with neither confirmed nor presumed diagnosis of ASD. Predictor: AQ (Autism Spectrum Quotient); outcome: BDI (Beck Depression Inventory); mediators: AAS (anx) = Adult Attachment Scale: attachment anxiety; AAS (avoid) = Adult Attachment Scale: attachment avoidance; MZQ = Mentalization Questionnaire; MSPSS = Multidimensional Scale of Perceived Social Support; AAQ-II = Acceptance and Action Questionnaire-II.

Group comparisons of indirect effects using Wald chi-square tests revealed that, in most cases, the size of the mediation effects did not significantly differ between groups (all *p-values* > .05). An exception was found for psychological inflexibility (AAQ-II), where the indirect effect was significantly larger in the non-ASD group compared to the ASD group (χ²(1) = 5.90, *p* = .015). The avoidant attachment pathway (AAS-avoid) remained stronger in the ASD group; however, after adjusting for covariates, the group difference was no longer statistically significant, though it approached trend level (Wald χ² = 3.01, *p* = .083). No significant differences were observed between groups for the other mediators, including mentalization (MZQ), anxious attachment (AAS-anxious), and perceived social support (MSPSS) (all Wald χ²(1) tests, *p* > .05). For detailed descriptions of Wald test comparisons see Supplementary Material Table S4.

## Discussion

Our investigation employed between-group comparisons and a spectrum approach, utilizing a moderated mediation method to analyze the relationship between autistic traits and depressive symptom severity across three groups: individuals with a formal ASD diagnosis, self-diagnosed autistic individuals (ASD-sd), and neurotypical controls. We examined five potential mediators: mentalizing, anxious and avoidant attachment style, perceived social support, and psychological flexibility. The findings indicated that the strength and pattern of mediation varied across groups. Despite significant differences in all scores between groups, psychological flexibility emerged as the strongest and most consistent mediator regardless of diagnostic status, while mentalizing and avoidant attachment showed significant indirect effects in some groups but not others. Notably, the mediation was complete: once the mediators were included, the direct association between autistic traits and depressive symptoms was no longer significant, highlighting the central role of psychosocial and cognitive processes in mediating this relationship.

Depression in autism presents a notable burden, not solely due to its high frequency, but also because of challenges in its recognition and accurate diagnosis in clinical practice. The onset of symptoms may be atypical or misattributed to core features of autism itself, such as social withdrawal or reduced emotional expression (Chandrasekhar & Sikich, 2015). Our findings confirm that individuals with higher autistic traits, particularly those in the autism group, report more severe depressive symptoms than neurotypical controls. However, the psychological mechanisms linking these traits to depression differ in strength and character across groups. This suggests that while individuals with elevated autistic traits are broadly at increased risk for depression, as also found in previous studies (Hollocks et al., 2019; Lai et al., 2019; Wigham et al., 2017), the pathways through which this risk manifests may vary by diagnostic group, underscoring the importance of identifying group-specific mechanisms to inform more tailored interventions.

One of the most notable findings of our study was that psychological flexibility was the strongest and most consistent mediator between autistic traits and depressive symptoms across all groups. This construct, defined as the ability to stay present while adapting behavior in line with personal values (Kashdan & Rottenberg, 2010), has been validated for use in autistic adults (Aller et al., 2022) and has been shown to buffer against emotional distress in both clinical and non-clinical populations. While autism is characterized by behavioral inflexibility (APA, 2022), psychological flexibility remains under-studied in this population. Previous research in neurotypical samples has consistently linked lower flexibility to higher levels of depression and maladaptive coping (Bond et al., 2011). A recent study by Lee et al. (2022) found that psychological flexibility, as measured by the AAQ-II, alongside attachment anxiety, mediated the association between adverse parenting experiences and later mental health outcomes in autistic individuals. This suggests that the capacity to adaptively respond to distressing internal states may be a crucial resilience factor across diagnostic boundaries. Consistent with this, theoretical models linking attachment and emotion regulation to psychological flexibility further support this idea (Mikulincer & Shaver, 2019; Zilberstein, 2014), highlighting overlapping pathways through which early relational experiences can shape later vulnerability. Despite being under-studied in autism, psychological flexibility appears to be a promising transdiagnostic construct with clear relevance for therapeutic interventions.

While psychological flexibility was the most universal pathway, other mediators, especially mentalization and avoidant attachment, showed group-specific effects. Mentalizing is often cited as a key factor in understanding depression in autism, in our study, it only emerged as a significant mediator in the non-ASD group. This result is somewhat unexpected given the extensive literature linking ASD with impairments in mentalizing and reflective functioning (Arioli et al., 2021; Brewer et al., 2017; Chung et al., 2014; Yirmiya et al., 1998). One possible explanation is that more severe or trait-like impairments in the ASD group may limit individuals’ ability to reflect on and accurately report their mentalizing capacity, a phenomenon that is paradoxically consistent with the very definition of compromised reflective functioning. In contrast, subclinical or milder deficits in the non-ASD group may remain accessible to introspection and thus detectable via self-report tools. This divergence highlights a limitation of self-report-based measurement of mentalizing (Wendt et al., 2024) and suggests that more nuanced, performance-based or informant-rated approaches might be necessary to capture its true role in this population. While mentalizing deficits in ASD are well-established, and comparable in severity to those observed in schizophrenia (Bliksted et al., 2016; Fernandes et al., 2018; Oliver et al., 2021), future studies should explore the dimensional profile of mentalization (e.g., cognitive vs. affective, self vs. other) using more sensitive tools, particularly in self-diagnosed populations, where mixed patterns may reflect distinct compensatory or coping strategies.

Avoidant attachment was the only variable that significantly distinguished the ASD and ASD-sd groups in terms of mean scores, with self-diagnosed individuals reporting higher levels than those with a formal diagnosis. Interestingly, however, its mediating effect between autistic traits and depressive symptoms was significant only in the clinically diagnosed ASD group. This contrast raises important questions about the mechanisms underlying depressive vulnerability in self-diagnosed individuals. This divergence suggests that while avoidant tendencies may be more frequently endorsed by self-diagnosed individuals, potentially due to prolonged unmet support needs or social exclusion (Camus et al., 2024; Overton et al., 2024), they may carry greater psychological weight and reflect deeper relational disengagement among formally diagnosed individuals. In the ASD group, avoidant attachment may reflect longstanding relational disengagement, potentially rooted in early social-emotional developmental differences. This aligns with previous studies showing a high prevalence of insecure attachment in autism (Rutgers et al., 2004; Van IJzendoorn et al., 2007) and its association with affect regulation difficulties (Dagan et al., 2018; Mikulincer & Shaver, 2019).

In contrast, in the ASD-sd group, higher avoidant scores may not translate as directly into depressive symptoms, possibly due to different meaning-making (attributional) processes or identity-related struggles that dominate their psychosocial experience (Davidson & and Henderson, 2010; Lewis, 2017). Instead their depressive symptoms could rather stem from social invalidation, lack of recognition, or difficulty accessing identity-affirming communities (Camus et al., 2024; Cooper et al., 2023). These individuals often report a weaker sense of belonging to the autistic community (Camus et al., 2024), which may reduce the protective effect commonly associated with community identification and support (Cooper et al., 2023; Crompton et al., 2024). Thus, while avoidant attachment is elevated in this group, its explanatory power for depression may be limited compared to that of other psychosocial mechanisms such as perceived isolation or unacknowledged identity. The role of attachment insecurity as a contributing, rather than central, mechanism may suggest indirect clinical relevance. Although this is not a primary therapeutic target, the patient’s attachment style could influence treatment engagement and emotional availability during therapy (Fischer-Kern et al., 2022; Reiner et al., 2016).

The observed pattern of indirect effects indicates that high AQ scores alone do not explain the emergence of depression, but rather their interaction with these cognitive-affective mechanisms. These findings are consistent with previous research on neurotypical populations with high autistic traits, who also report more depressive symptoms when mentalizing capacity is impaired or when support is perceived as lacking (Beatson & Taryan, 2003; Besser & Priel, 2003; Carnelley et al., 1994; Malik et al., 2015). The overlap between attachment insecurity, poor mentalizing, and limited psychological flexibility may also hinder individuals in expressing their needs or seeking help— factors that are particularly relevant for those without formal diagnosis and who may not access support services (Davidson & and Henderson, 2010; Lewis, 2017).

Finally, perceived social support, while not the strongest mediator overall, remained an important indirect factor in the non-ASD group. Earlier findings showed that perceived lack of support, combined with social anxiety and camouflaging behaviors, contributes to depressive symptoms (Bernardin et al., 2021; Hull et al., 2017; Lai et al., 2019). For self-diagnosed individuals, these effects may be exacerbated by uncertainty around identity and lower community integration, as suggested in recent studies (Camus et al., 2024; Overton et al., 2024). This supports the need to conceptualize self-diagnosed autism not merely as a milder form of clinical ASD, but as a distinct presentation that may entail equal or greater emotional burden, especially in relation to unacknowledged needs and difficulties accessing appropriate support.

### Clinical relevance and therapeutic implications

A significant number of individuals with autism spectrum disorder seek therapy for depression, presenting unique challenges for effective treatment. Managing (both recognising and treating) depression in this population is particularly difficult due to the complex interplay of social, cognitive, and emotional factors inherent to ASD. Our findings underscore the need for personalized, mechanism-informed interventions for depression in individuals with elevated autistic traits. Rather than targeting autistic traits directly, therapeutic efforts may be more effective when focused on transdiagnostic cognitive-affective processes, particularly psychological flexibility, mentalizing capacities, and attachment-related functioning.

Psychological flexibility emerged as the most robust and universal mediator of depressive symptoms across all groups, highlighting its value as a core resilience factor and key intervention target. Enhancing flexibility could reduce emotional rigidity and support adaptive functioning, particularly through structured approaches such as acceptance and commitment therapy, which has growing empirical support in both neurotypical and autistic populations (Aller et al., 2022; Pahnke et al., 2023).

Avoidant attachment, while more prevalent in the self-diagnosed group, exerted its mediating effect only in the clinically diagnosed ASD group, suggesting that avoidant interpersonal strategies may carry different meanings and consequences across diagnostic contexts. From a therapeutic perspective, high levels of avoidant attachment can undermine treatment engagement, emotional attunement, and the therapeutic alliance. Identifying these patterns early and tailoring relational dynamics may improve adherence and outcomes (Fischer-Kern et al., 2022; Reiner et al., 2016). Clinical experience drawn from mentalization-based treatment (MBT) protocols for avoidant personality disorder suggests that patients with pronounced relational withdrawal benefit from carefully paced interventions that scaffold mentalizing at a comfortable pace (Bateman et al., 2023). These strategies emphasize trust-building and indirect engagement and may be highly applicable to autistic individuals around avoidant attachment, supporting earlier identification and personalized pacing in therapy.

Although mentalizing did not mediate depressive symptoms in the ASD group, this may reflect the limits of self-awareness in this population, rather than a lack of clinical relevance. Mentalization-based treatment, with its focus on enhancing reflective functioning in emotionally charged contexts, may offer a promising yet underutilized avenue, provided that it is adapted to the cognitive and social profiles of autistic individuals. Recent efforts to tailor MBT to ASD contexts, while still limited, show preliminary promise (Cassel et al., 2007; Krämer et al., 2021).

Finally, the findings emphasize the importance of context-sensitive care for self-diagnosed individuals, who often do not fit into existing diagnostic frameworks but nevertheless face clinically significant distress. Interventions for this group should consider issues of identity, belonging, and social validation, which may lie at the core of their depressive vulnerability.

In summary, our results support a shift away from diagnosis-based assumptions and toward process-based treatment models, aligning with contemporary calls for personalized, transdiagnostic care in both autism and mental health services more broadly.

### Limitations

A major strength of our study is the use of self-report questionnaires administered online, which enabled the collection of data from a relatively large and diverse sample. However, several limitations must be acknowledged. First, the validity of self-report measures may be influenced by participants’ introspective accuracy, particularly for constructs like mentalizing, where limited self-reflection may bias responses. Nonetheless, previous research suggests that subjective perceptions may be more predictive of psychopathology than objective adversity alone (Danese & Widom, 2020).

Second, while the ASD group included individuals with formal clinical diagnoses, the ASD-sd group was composed of self-identified autistic individuals without diagnostic confirmation. Although we analyzed them as a distinct group, the heterogeneity within the ASD-sd population remains a potential limitation, given that motivations for self-identification and levels of self-awareness may vary widely. Furthermore, it cannot be ruled out that undiagnosed autistic individuals may be present in the non-ASD group, potentially diluting group contrasts.

Additionally, the online recruitment strategy may have introduced sampling bias, favoring individuals who are more self-aware, better resourced, or more engaged with autism-related content. Importantly, we did not assess the effect of comorbid psychiatric diagnoses or medication use, both of which may influence depressive symptoms and the mechanisms we examined. Although gender was included as a covariate in our models, the overall sample showed a gender imbalance, limiting the generalizability of our findings and highlighting the need for future studies with more representative and stratified samples.

Finally, the cross-sectional design of our study precludes causal interpretations. Longitudinal studies are needed to assess the temporal dynamics between autistic traits, mediating psychosocial factors, and depressive symptoms.

## Conclusions

This study provides a nuanced understanding of how psychosocial and cognitive factors mediate the link between autistic traits and depressive symptoms across formally diagnosed, self-identified, and neurotypical populations. Among the mediators examined, psychological flexibility emerged as the most robust and consistent protective factor, while avoidant attachment and mentalizing difficulties showed group-specific relevance. These findings highlight the need for targeted, mechanism-based interventions that move beyond categorical diagnoses. Enhancing psychological flexibility and addressing relational patterns, especially avoidant attachment, may be particularly effective therapeutic entry points. Mentalization-based treatment, when appropriately adapted, holds promise as a framework that integrates these mechanisms. Ultimately, our results underscore the value of personalized approaches that recognize the heterogeneity of autistic experiences and prioritize transdiagnostic resilience-building strategies in the prevention and treatment of depression.

## Data availability statement

The data that support the findings of this study are available on request from the corresponding author.

## Ethics approval statement

This study was approved by the Semmelweis University Regional and Institutional Committee of Science and Research Ethics, approval number SE RKEB: 159/2021.

## Patient consent statement

Informed consent was obtained from all participants for this study. Participants indicated their consent to participate by clicking the provided button after reading the informed consent form.

## Permission to reproduce material from other sources

Not applicable.

## Clinical trial registration

Not applicable

